# Characterizing Two Outbreak Waves of COVID-19 in Spain Using Phenomenological Epidemic Modelling

**DOI:** 10.1101/2020.12.29.20247833

**Authors:** Miguel López, Alberto Peinado, Andrés Ortiz

**Affiliations:** Universidad de Málaga, Andalucía Tech – BioSip Research Group, E.T.S. Ingeniería de Telecomunicación, Dept. Ingeniería de Comunicaciones, Campus de Teatinos 29071, Málaga, Spain

**Author notes:** {, }.

**Keywords:** COVID-19, Epidemics, Spain Incidence, Phenomenological modelling

## Abstract

Since the first case reported of SARS-CoV-2 the end of December 2019 in China, the number of cases quickly climbed following an exponential growth trend, demonstrating that a *global pandemic* is possible. As of December 3, 2020, the total number of cases reported are around 65,527,000 contagions worldwide, and 1,524,000 deaths affecting 218 countries and territories. In this scenario, Spain is one of the countries that has suffered in a hard way, the ongoing epidemic caused by the novel coronavirus SARS-CoV-2, namely COVID-19 disease. In this paper, we present the utilization of phenomenological epidemic models to characterize the two first outbreak waves of COVID-19 in Spain. The study is driven using a two-step phenomenological epidemic approach. First, we use a simple generalized growth model to fit the main parameters at the early epidemic phase; later, we apply our previous finding over a logistic growth model to that characterize both waves completely. The results show that even in the absence of accurate data series, it is possible to characterize the curves of case incidence, and even construct short-term forecast in the near time horizon.

## 1. Introduction

An infectious disease is characterized because it can be transmitted, either directly or indirectly, from one individual to another within a given population. This transmission is materialized in an increase in the number of newly infected individuals per unit time, representing the infectious disease incidence. For example, an infected individual may remain asymptomatic at the early stage of infection, only later developing clinical symptoms, being diagnosed as a disease case. If the number of cases rises above the usual average within a short period of time, a *disease outbreak* occurs, but if the disease spreads quickly to many people, it is an *epidemic*. When the disease persists, and may remain in the population over a long period of time, the disease is said to be *endemic* in the population. Finally, if the disease spreads spatially on a global scale to many countries and continents, a *global pandemic* occurs [1].

At the end of December 2019, a set of pneumonia cases arisen in the city of Wuhan (Hubei province, China). On January 7, 2020, the Chinese authorities notified a new virus from the Coronavirus family as the probably causative agent of the outbreak namely SARS-CoV-2. The disease caused by this new virus has been called by international consensus COVID-19, and the World Health Organization (WHO) recognized it as a global pandemic on 11 March 2020 [2]. As of December 3, 2020, the total number of COVID-19 the reported cases are around 65,527,000 contagions worldwide, and 1,524,000 deaths affecting 218 countries and territories.

Nowadays, the ongoing epidemic caused by the novel coronavirus SARS-CoV-2, represents the more recent global pandemic arisen in decades; demonstrating that a *global pandemic* is possible, showing its potential to generate explosive outbreaks not only in cities or regions, but also in countries or continents following human mobility patterns [4]. While the symptoms observed in COVID-19 infected people are frequently mild, and quite similar to other common respiratory infections, it has also exhibited an ability to generate severe disease among certain groups, including older populations and individuals with underlying health issues such as cardiovascular disease and diabetes [5]. Nevertheless, the transmission patterns and other epidemiology characteristics of this novel coronavirus are still being studied.

In this scenario, Spain is one of the countries that has suffered the COVID-19 pandemic in a hard way. The presence of the virus was first confirmed in Spain on January 31, 2020. Since then, the number of cases quickly climbed following an exponential growth trend, reaching all provinces at March 13, 2020. At March 14, 2020, the Spanish Government imposed a confinement or lockdown that was maintained until June 21, 2020 [3, 7]. This first epidemic outbreak wave, reached its peak at March 20 with 10,651 cases reported and 996 deaths at April 2. The total cumulative number of cases as of June 21, is of 255,547, and 29,353 deaths [8]. After the end of the confinement, it was observed a period of stability in the incidence of the disease accounting for a low number of reported daily cases around the country. However, the number of new cases starts climbed in July in various cities, presenting again an exponential growth trend profile, which has remained during the last months. As of December 3, 2020, the total number of COVID-19 the total cumulative number of cases accounts around 1,693,600 contagions, including 46,000 deaths [3]. This is hence, the so called second wave which is currently ongoing.

This scenario drive epidemiologist, mathematicians and other scientist to study and forecast the expected behavior of the epidemic outbreak and its dynamics. These studies are intended to assess public health authorities to take appropriate decisions about control measures strategies, in order to mitigate the impact of the disease in the absence of an effective vaccine.

In this paper, we present the utilization of phenomenological epidemic models to characterize the two first outbreak waves of COVID-19 in Spain. The study is driven using a two-step approach to fit the main parameters that characterize both waves. First, a simple growth model is used to identify the intrinsic growth rate and the deceleration parameter for the first and the second wave, during the early stage of the epidemic. Further, these parameters are applied in a second step using a logistic growth model, obtaining a complete characterization of the first wave, and a forecast of the behavior of the second wave. Our main goal is, through simulations and empirical data, to understand and characterize the current state of the epidemics waves outbreaks of the COVID-19 in Spain. Also, we estimate the possible pandemic cessation dates, and the total size of the epidemics at the end of the second wav by constructing mean-time forecasting e. Hence, this study could be regarded as a valuable tool for characterizing the transmission dynamics process of COVID-19 pandemic along with the impact of control measures, when they were fully implemented and sustained.

The rest of the paper is organized as follows. In Section 2 introduces the basics of phenomenological epidemic models. Section 3 presents the parameter estimation process. Simulation study based on the reported case incidence is presented in Section 4. Section 5 gives the results and discussion; and finally, future works are proposed in Section 6.

## 2. Basics of phenomenological epidemic models

Classical epidemic models are based on mechanistic or physical approaches in order to identify patterns in the observed data, comprising the laws involved in the dynamics of the population or transmission dynamics of a disease. These models such as Kermackand–McKendrick [9] or Anderson and May [10] are based on compartments of population in which it is assumed that in the absence of control strategies, the growth dynamics in the first stage of an epidemic is exponential. By contrast, phenomenological epidemic models provide an empirical approach without a specific basis on the physical laws or mechanisms that give rise to the observed patterns in the data. Most of commonly used phenomenological epidemic approaches are based on population growth models [18].

### 2.1. The Generalized Growth Model (GGM)

The Generalized Growth Model (GGM) is based on phenomenological population growth dynamics. The model emphasizes in the reproducibility of empirical observations and have been proven to be very useful to model early epidemics in various diseases such as SARS, Ebola, Zika or COVID-19 [11, 12, 13, 14, 15]. This model assumes sub-exponential growth dynamic at the beginning of the disease rather than exponential. The GGM relies on two parameters to relax the assumption of exponential growth and is defined by the differential equation 1 as follows [16]:

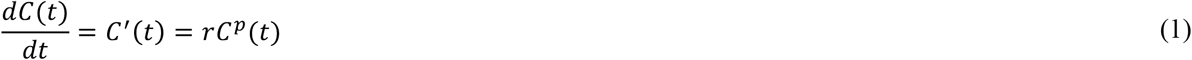

Here, *C*′ (*t*) represents the incidence at time *t*, whereas the solution *C*(*t*) is the cumulative number of cases at time *t*. The parameter *r* is a positive value denoting the intrinsic growth rate (1/*t*), and the parameter *p* is considered the *deceleration of growth* factor where *p* ∈ [0, 1]. If *p* = 0 this differential equation describes a constant incidence curve over time, where the cumulative number of cases grows linearly. By contrast, if *p* = 1 it models exponential growth dynamics, and the solution of the equation is *C* (*t*) = *C*_0_*e*^*rt*^, where *C*_0_ is the initial number of cases. Choosing intermediate values of *p* whiting 0 and 1, the equation describes sub-exponential growth behavior. For example, for *p* = 1/2 incidence grows linearly while the cumulative number of cases follows a quadratic polynomial. For *p* = 2/3 incidence grows quadratically while the cumulative number of cases fits a cubic polynomial [11]. This model can also support different epidemic growth patterns depending on the interval in which the value of *p* moves. This includes linear incidence patterns for *p* = 0.5, concave-up incidence patterns for 0 > *p* > 0.5, and concave-down incidence patterns for 0.5 < *p* < 1 [11].

### 2.2. The Generalized Logistic Growth Model (GLGM)

Despite GGM provides an adequate approximation to for the initial phase or *early epidemics* of the transmission dynamics of a disease, this model does not take into account reductions in the disease incidence due to the characteristics of the pathogen, the immunity of the population, or the implementation of medical or social measures. Therefore, consider unlimited growth of contagion is quite unrealistic. To solve this issue, some epidemic models considers logistic growth, in which for a given stable population, it would have a saturation level which representing a numerical *upper bound* for the growth size of the epidemic. This bound is typically called the *carrying capacity K* and in this model, represents the final size of the epidemic. This parameter is crucial to estimate how severe could be the disease, since it represents the final size of the population that became infected. The GLGM is defined by the differential equation 2 [17, 18]:

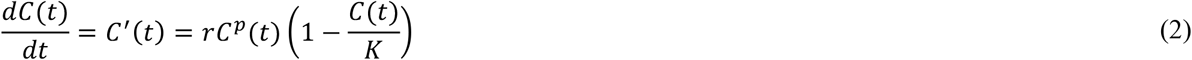

where *C(t)* represents the cumulative number of cases at time *t, r* is the intrinsic growth rate, *p* is the scaling of growth parameter. As in the GGM *p* = 1 indicates early exponential growth, whereas *p* = 0 represents constant growth, and 0 < *p <* 1 accommodates early sub-exponential or polynomial. *K >* 0 is the carrying capacity or final size of the epidemic, which represents the total number of affected population. This parameter is crucial to generate post-peak forecasts and also, can be linked to an import parameter of classical epidemic models such as the basic reproduction number *ℛ*_0_, which represent the transition phase of non–equilibrium process of propagation of a disease. This parameter constitutes a key epidemiological threshold in biological systems since if *ℛ*_0_<1 the infection dies out while if *ℛ*_0_>1 it may cause an epidemic disease. In fact, assuming that COVID-19 can be modelled as the simplest but useful SIR (*Susceptible-Infected-Recovered*) epidemic model [1], the basic reproductive number can be calculated as *ℛ*_0_ = *S*_0_/*ρ*, where *ρ* = (*S*_0_–*S*_∞_)/(*ln S*_0_–*ln S*_∞_), *S*_0_ is the initial population of susceptible, and *K* = *S*_0_–*S*_∞_ is the final size of the epidemic.

## 3. Parameter estimation

Parameter estimation can be considered as the process of finding parameter values and their confidence intervals that best fit a given model, to the empirical data. In this study, we follow the methodology presented in [14, 19, 20] that has been used to characterize the incidence of various infectious diseases such as Ebola, Smallpox, Measles, or Zika. In essence, this methodology comprises a process that starts with the initial adjustment of the data, then the parameters for each model are fitted based on early epidemic phase. Since these parameter estimations are typically subject to sources of uncertainty, it is constructed the 95% confidence intervals derived from parameter uncertainty.

### 3.1. Data sources

Since the beginning of the pandemic in Spain, surveillance of cases of this disease is based on the universal notification of all confirmed COVID-19 cases that are identified in each Autonomous Community [7]. The Autonomous Communities (CCAA) make an individualized notification of COVID-19 cases through a web-based computer platform managed by the National Epidemiology Center (CNE). This information comes from the epidemiological case survey that each Autonomous Communities completes upon identification of a COVID-19 case. Despite the fact that some studies have suggested that the number of infections and deaths in Spain may have been underestimated [21], for simulation purposes our study is based on the case incidence reported by the Ministry of Health of Spain, and brought as *csv* file in the official data repository [7]. The time interval comprises 44 weeks (308) days, ranging from January 31, 2020, to December 3, 2020. Figure 1, shows the profile of daily cases reported as blue circles (Fig. 1.a), as well as the curve of cumulative number of cases blue solid line (Fig. 1.b). In both pictures, it is easy to identify the two epidemics waves.

**Figure 1.**
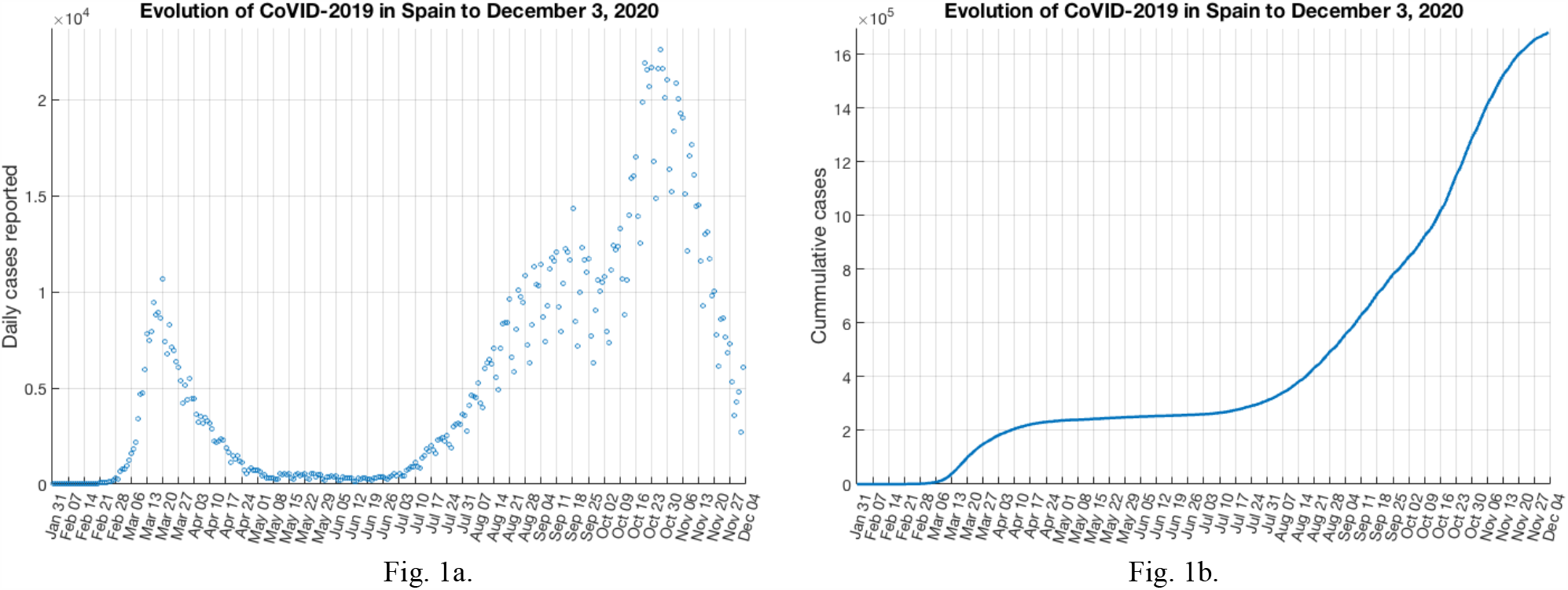
Profile of daily cases reported (Fig. 1.a), and the curve of cumulative number of cases (Fig. 1.b) in Spain from January 31, 2020, to December 3, 2020.

### 3.2. Parameter fitting

One of the simplest way to estimate the parameters from observed data is using nonlinear least squares fitting. This method tries to obtain the solution that best-fit the curve of the desired model, given a set of data points. For example, aplying a non-linear least-squares solver, that uses a *Trust-Region-Reflective* algorithm (or the *Levenberg–Marquardt* algorithm) to provide model coefficients that minimize the sum of squared differences between the model and a set of data [22]. Basically, these methods considers a parameter set *θ* = (*θ*_3_, *θ*_5_, …, *θ*_*k*_), the family of curves *y* = *f*(*t, θ*) that depend on parameter *θ*, and a set of data points (*t*_1_, *y*_1_), (*t*_2_, *y*_2_), …, (*t*_*n*_, *y*_*n*_). The nonlinear least-squares fitting, tries to find the value of coefficients 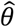 such that the curve 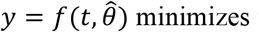 minimizes the objective function:

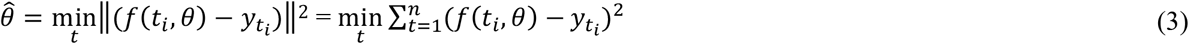

In order to use a least-squares fitting correctly, it should be defined for the desired parameters *θ*, the upper and lower bounds, and an initial guess. In the case of GGM *θ* = (*r, p*) and for GLGM *θ* = (*r, p, K*). Also, after the parameters have been estimated, the function provides the residuals, as the difference between the best fit of the model and the time series data as a function of time, in the form 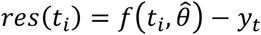. This is used to assess the quality of the model fit.

### 3.2. Quantifying parameter confidence intervals

Before the generation of the short-term incidence forecast we still need to quantify the uncertainty of the parameter estimates to construct the 95% confidence intervals, and to identify the potential parameter deviation. One of the methods to quantify parameter uncertainty is the *parametric bootstrapping* [23, 24]. Basically, this method re-estimates the parameters from the previously best-fit model 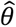, by sampling repeatedly multiple observations and generating synthetic datasets. Also, it is considered that the time series follow some distribution error structure (binomial, Poisson, etc.). This method can be implemented in different ways [19, 24]. For this study, we use the parametric bootstrap approach proposed in [19] that has been applied to model various infectious diseases such as Ebola, Smallpox, or Zika [14, 15]. This method assumes that the time series of data follows a Poisson distribution error structure, centered on the mean of the time points of each observation [25]. For a given set of parameter 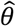, a parametric bootstrapping of *m* realizations it is derived a new set of re-estimated parameters 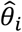, where *i* = 1, 2,…, *m*. The resulting uncertainty around the new model fit is given by the set of curves 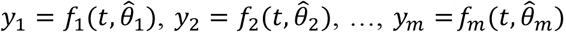.

### 3.4. Short-term forecasting of incidence curves

The previous calibration of GGM parameters, together the confidence intervals provides a characterization of the early epidemics respect the first and second wave. But we also aim to characterize the profile of the first epidemic wave, and the expected behavior of the second epidemic wave in the near time horizon. In the case of the COVID-19 pandemic, short-term forecast (days or weeks) is useful to develop medical plans or to schedule contention measures in the absence of an effective vaccine. However, it is important to keep in mind that forecasts are often inaccurate as these are mostly based on the current values and uncertainty of the parameters of the system, which are likely to change over time. Moreover, the further out the forecast is made, the more wrong it is expected to be [19]. One method to construct short-term forecast, is to extend the entire uncertainty of the system using the uncertainty associated to the previously computed parameter estimates [19]. In this way, the current state of the system in a given time, is propagated to a time horizon of *T* time units ahead. Then, the set of curves 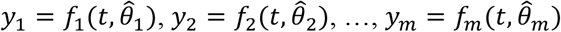 are newly computed as 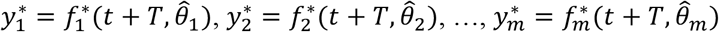.

## 4. Simulation study

### 4.1. Characterizing the early epidemic phase

In this simulation, we start estimating the parameters *r* and *p* for the first and second wave using the GGM approach. For the first wave, it is assumed an early epidemic period of 38 days, (from February 6, 2020 to March 14, 2020), and the initial number of cases reported *C*_0_ = 11. For the second wave, it is assumed an early epidemic period of 45 days (from, July 30, 2020, to August 14, 2020) and the initial number of cases reported *C*_0_ = 540. To compute the least-squares fitting, the guess for parameters was set for both waves as *r* = 0.5 and *p* = 0.5, and the bounds as 0< *r* <10 and 0< *p* <1 respectively. Figure 2 shows for the GGM model, the best fit of the early epidemic period for the first wave (Fig. 2.a) and for the second wave (Fig. 2.b) of the COVID-19 in Spain. The blue circles represent the reported daily case incidence, while the continuous red line corresponds to the best fit of the GGM to the data. Also, it has been displayed the residuals 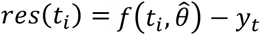. in the right panel of each figure. It should be noted the random pattern of the residuals as a function of time, which suggest that the GGM model has provided a reasonably good fit to the early growth phase of the epidemic.

**Figure 2.**
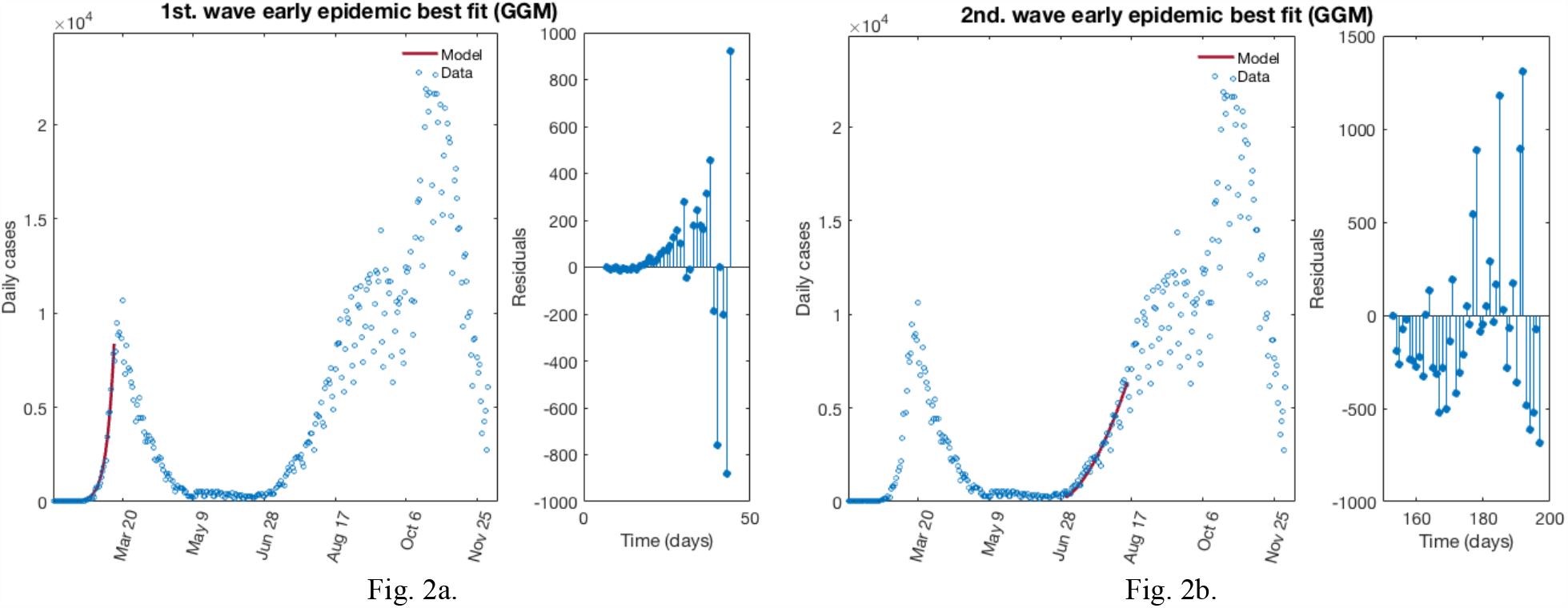
The best fit for GGM of the early epidemic period for the first wave (Fig. 2.a) and second wave (Fig. 2.b) for COVID-19 in Spain. Blue circles represent the reported daily case incidence, while continuous red line corresponds to the best fit of the model. The residuals are displayed in the right panel each wave.

After determining the best fit for *r* and *p*, now we build the 95% confidence intervals by quantifying the parameter uncertainty. Figure 3 displays the re-fitting of parameters of the early epidemic phase for the first and the second wave using the GGM. The model has been calibrated using *m* = 300 bootstrap realizations, and assuming a Poisson distribution error structure. The blue circles are the cases reported daily, while the solid red line corresponds to the best fit of the GGM to the data. The grey lines correspond to the *m* bootstrapping realizations of the epidemic curves. The black lines correspond to the 95% of confidence interval (CI) bounds around the best fit of the model to the data. The top panels show histograms displaying the empirical distributions for parameter estimations.

**Figure 3.**
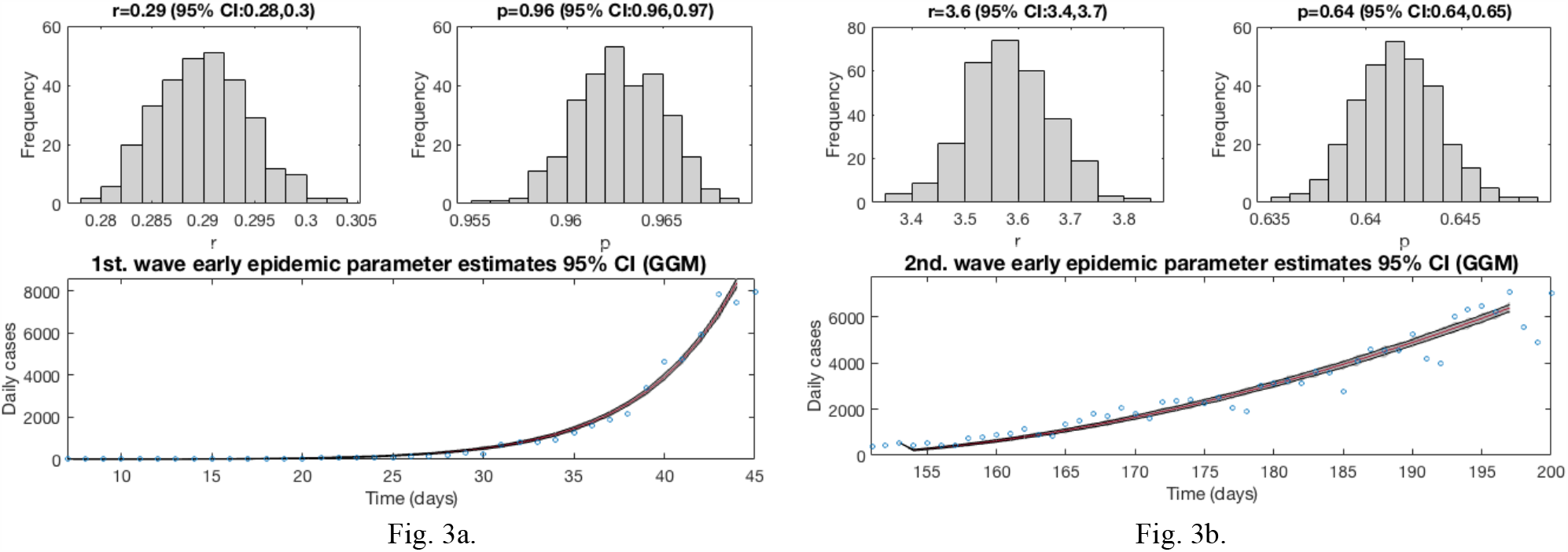
The best fit for GGM of the early epidemic period for the first wave (Fig. 3.a) and second wave (Fig. 3.b) for COVID-19 in Spain using 300 bootstrap realizations. Blue circles are the cases reported daily; red line corresponds to the best fit of the GGM to the data. Grey lines correspond to the *m* bootstrapping realizations of the epidemic curves. Black lines correspond to the 95% CI bounds. The empirical distributions for parameter estimations are displayed in the top panel for each wave.

Also, it has been constructed a short-term forecast of 7 days for the first wave (from, March 14, 2020 to March 20, 2020), and of 21 days for the second one (from August 14, 2020, to September 3, 2020), by extending the entire uncertainty from the early epidemic period, as described in section 3.4. Figure 4 shows the resulting curves. The calibration period is separated from the forecast period with a vertical slashed grey line. All the curves at the right side of this line, correspond to the forecast time horizon. The red line corresponds to the best fit of the forecasted epidemic curve. The black lines correspond to the 95% confidence bounds around the best fit of the forecasted curves, and grey lines correspond to the *m* bootstrapping realizations of the forecasted epidemic curves.

**Figure 4.**
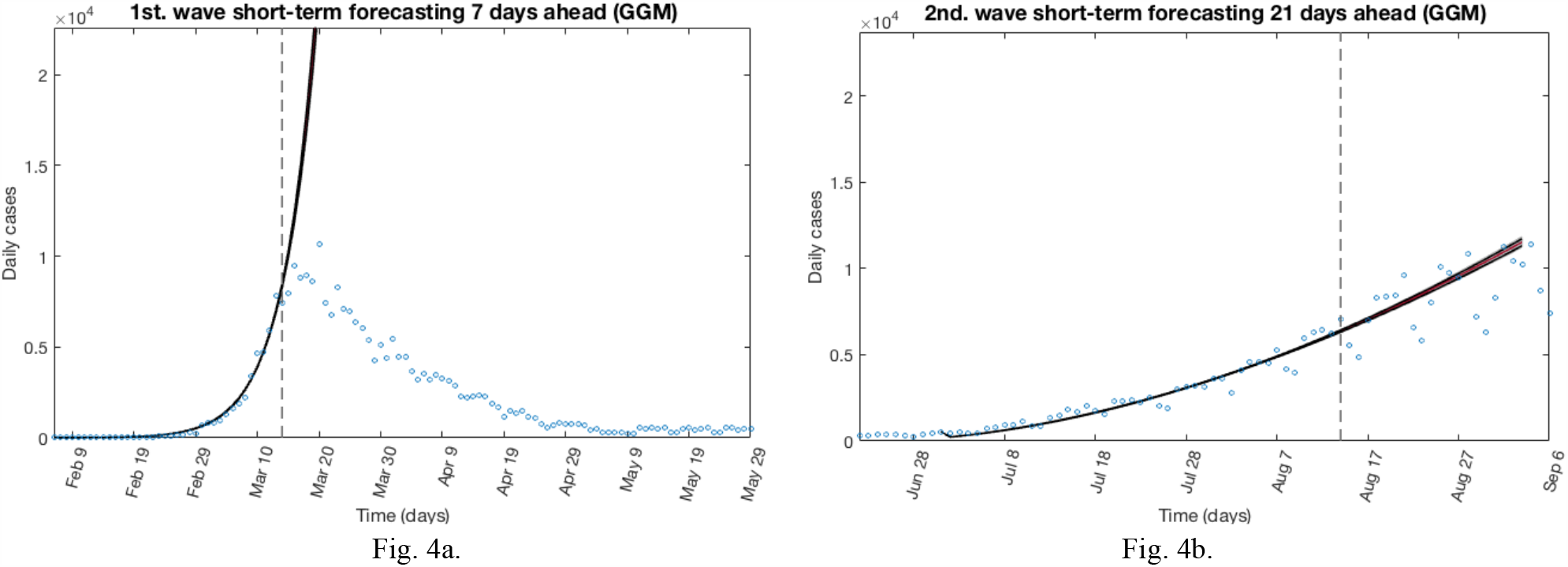
Forecasted curves for GGM for the first wave (Fig. 4.a) and second wave (Fig. 4.b) using 300 bootstrap realizations. The red line corresponds to the best fit of the forecasted epidemic curve. The black lines correspond to the 95% confidence bounds around the best fit of the forecasted curves, and grey lines correspond to the *m* bootstrapping realizations of the forecasted epidemic curves. The calibration period is separated from the forecast period with a vertical slashed grey line.

### 4.2. Characterizing the complete epidemic waves dynamics

The previous calibration of GGM and the resulting confidence intervals provide a properly adjustment model to data of the early epidemic for the first and second wave. However, GGM only considers unlimited growth of contagion, thus it is not adequate to model an epidemic that has entered or surpassed the peak of contagions. By contrary, GLGM includes the *upper bound* for the growth size of the epidemic to better characterize the after-peak dynamics of the epidemics, but brings poor adjustment to data in the early stage of the epidemic. Hence, to solve both problems, for the early stage of the epidemic of GLGM we will use the previous parameters and the resulting confidence intervals computed for GGM. Later, assuming that at the time to write this article, the first epidemic wave is finished and that the second wave is ongoing, but approaching to the peak of the maximum case reported, we use the GLGM to better characterize the whole dynamics of the epidemics.

To start with, now we estimate the best fit of the early epidemic using GLGM approach. It should be noted that GLGM is quite sensitive to initial definition of guess parameters as well as to the time intervals defined for the early epidemic phase. Therefore, previous to the definition of that values, we performed some simulations in order to better understand the dynamics of this epidemics. For the first wave, it is assumed an early period of 38 days, (from February 6, 2020 to March 14, 2020), and the initial number of cases reported *C*_0_ = 11. To compute the least-squares fitting, we use the mean of previous parameters estimation from GGM, thus the guess for parameters was set as *r* = 0.29 (95% CI: 0.28, 0.30) and *p* = 0.96 (95% CI: 0.96, 0.97), and the bounds was set as 0< *r* <10 and 0< *p* <1. Also, we stablish the limits for *K* at 1< *K* <47.3·10^6^, where the upper limit is the population of Spain at January 2020 [7]. Assuming that *K* is unknown, we estimate the final size of the epidemic as of 100,000 cases reported at the end of the first wave. For the second wave, it is assumed an early epidemic period of 55 days (from, July 8, 2020, to September 2, 2020) and the initial number of cases reported *C*_0_ = 898. Again, we use the mean of previous parameters estimation from GGM. Hence, the guess for parameters was set as *r* = 3.6 (95% CI: 3.40, 3.70) and *p* = 0.64 (95% CI: 0.64, 0.65), and the bounds was set as 0< *r* <10 and 0< *p* <1. The limits for *K* are stablished as 2.4·10^5^ < *K* <47.3·10^6^, where the lower limit is the mean value of the final size of the epidemic obtained from the calibration of the early epidemic of the first wave using GLGM. Since the second wave is still ongoing, *K* is unknown and we estimate the final size of that epidemic at 357,110 cases reported.

Now, after determining the best fit for both waves, we re-estimate the parameters *r, p* and *K* for the GLGM, and build the 95% confidence intervals by quantifying the parameter uncertainty. Figure 6 displays the re-fitting of parameters of the early epidemic phase for the first and the second wave using the GLGM. The model has been calibrated using *m* = 300 bootstrap realizations, and assuming a Poisson distribution error structure. The blue circles are the cases reported daily, while the solid red line corresponds to the best fit of the GLGM to the data. The grey lines correspond to the *m* bootstrapping realizations of the epidemic curves. The black lines correspond to the 95% of confidence interval (CI) bounds around the best fit of the model to the data. The top panels show histograms displaying the empirical distributions for parameter estimations.

**Figure 5.**
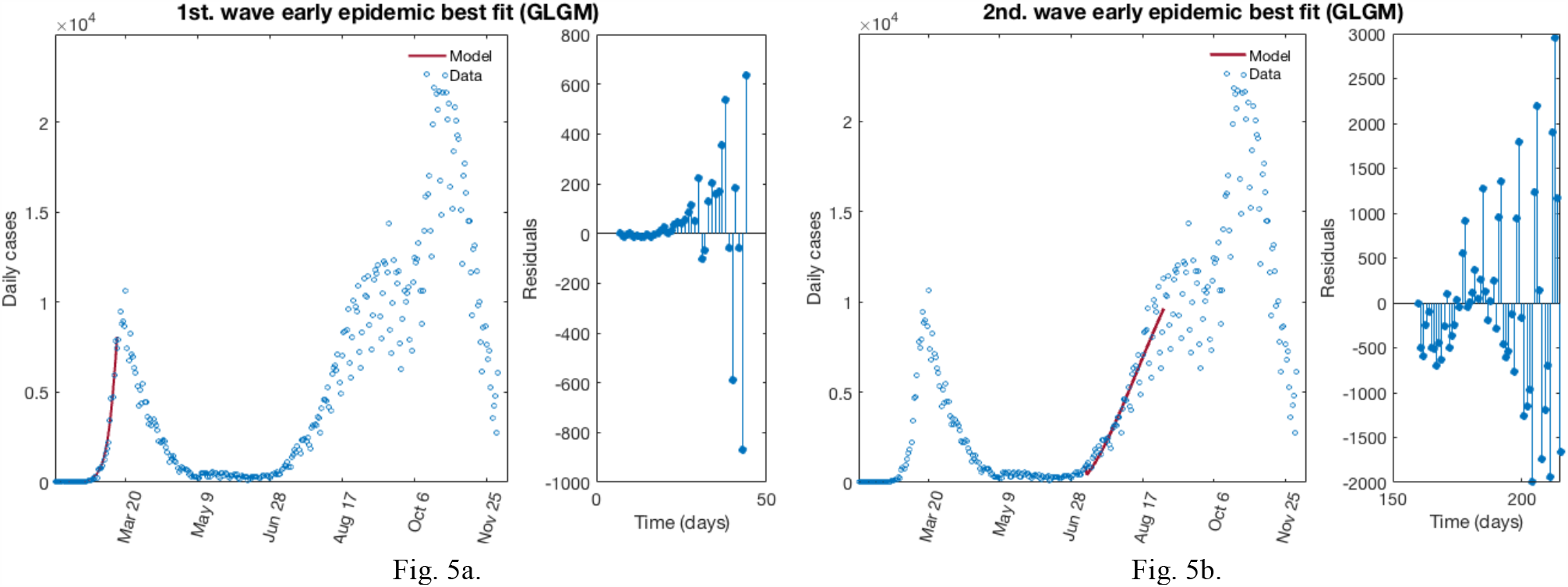
The best fit for GLGM of the early epidemic period for the first wave (Fig. 5.a) and second wave (Fig. 5.b) for COVID-19 in Spain. Blue circles represent the reported daily case incidence, while continuous red line corresponds to the best fit of the model. The residuals are displayed in the right panel each wave.

**Figure 6.**
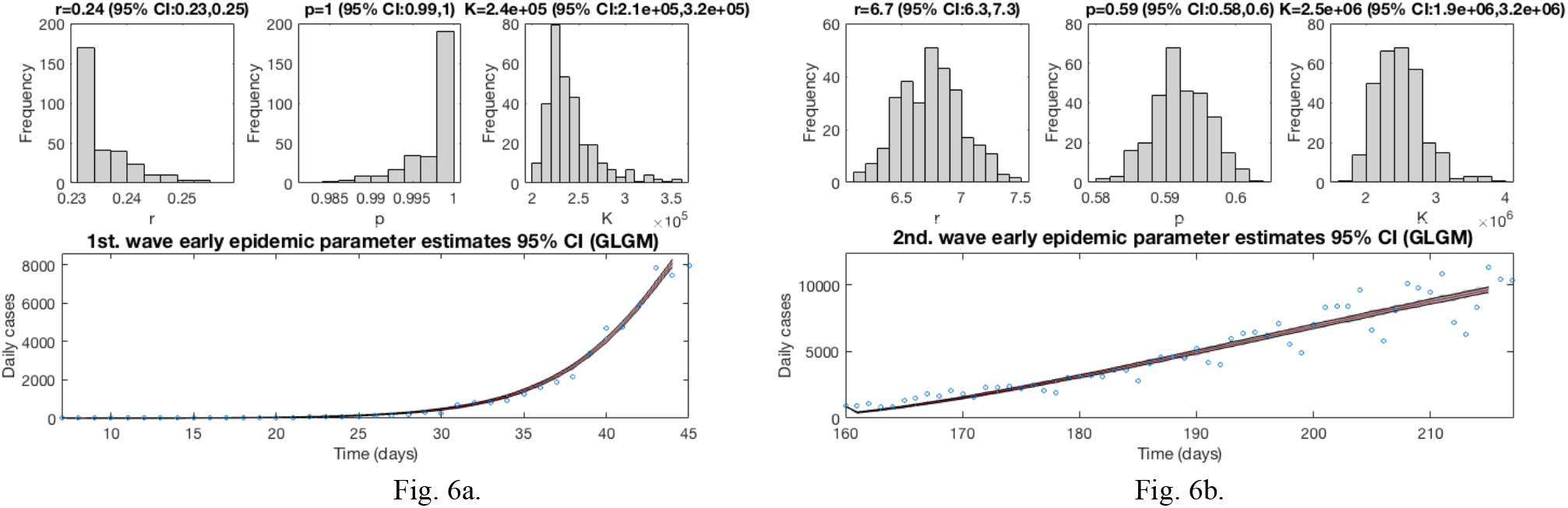
The best fit for GLGM of the early epidemic period for the first wave (Fig. 6.a) and second wave (Fig. 6.b) for COVID-19 in Spain using 300 bootstrap realizations. Blue circles are the cases reported daily; red line corresponds to the best fit of the GLGM to the data. Grey lines correspond to the *m* bootstrapping realizations of the epidemic curves. Black lines correspond to the 95% CI bounds. The empirical distributions for parameter estimations are displayed in the top panel for each wave.

Based on the previous calculations, now we can determine the epidemic profile for each wave by extending the entire uncertainty to a concrete period of time as described in section 3.4. For example, taking into account that the first wave is finished, we can add to the early epidemic period, a forecast of 108 days ahead to June 20, 2020. Figure 7 shows for the GLGM model, the best fit characterization of the first wave using *m* = 300 bootstrap realizations. In Figure 7.a, the blue circles are the daily cases reported. The solid grey lines correspond to the *m* bootstrapping realizations of the epidemic curves. The solid red line corresponds to the mean confidence or best fit of the GLGM respect the data; while solid black lines correspond to the upper and lower bounds of 95% confidence around the best fit of the model to the early epidemic. Also, it has been included in this figure, the curve of cumulative number of cases represented in Fig. 7b as the solid blue line; the red line corresponds to the best fit of the GLGM to the data. Grey lines correspond to the m bootstrapping realizations of the epidemic curves. Black lines correspond to the 95% CI bounds.

**Figure 7.**
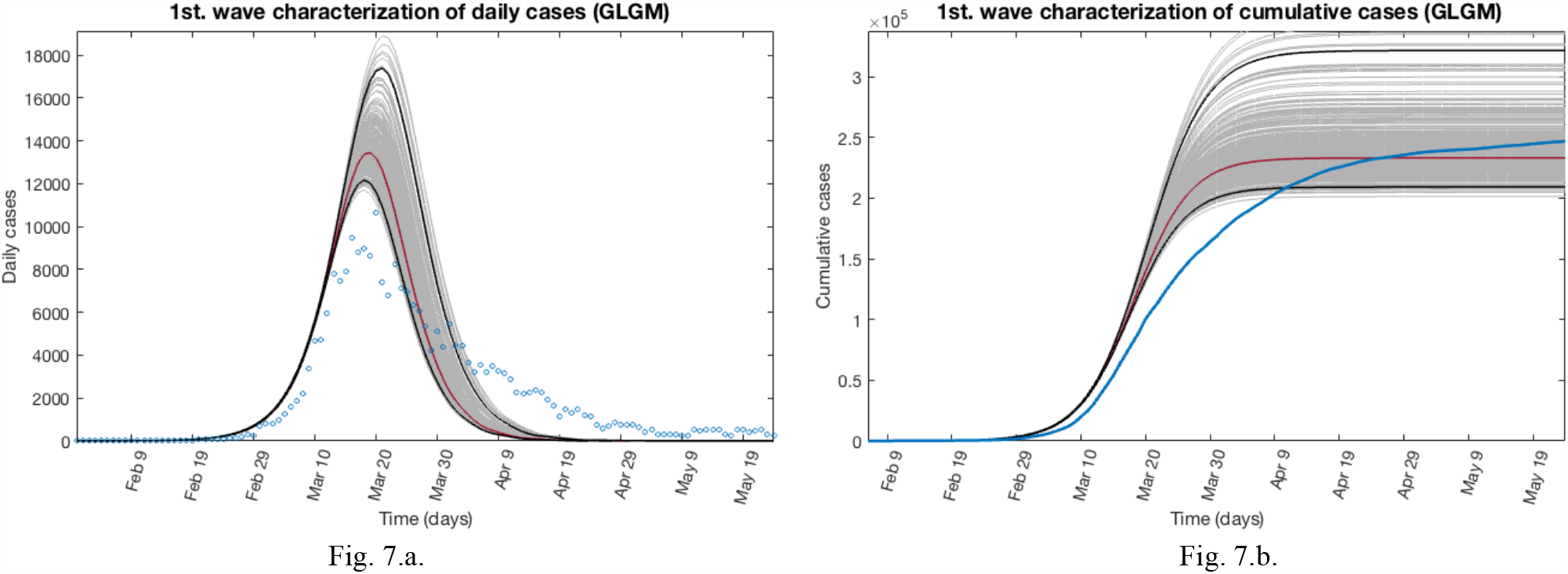
Characterization of the epidemic profile for the first wave using GLGM. In Fig. 7.a the blue circles are the cases reported daily; red line corresponds to the best fit of the GLGM to the data. Grey lines correspond to the *m* bootstrapping realizations of the epidemic curves. Black lines correspond to the 95% CI bounds. In Fig. 7.b, the solid blue line is the cumulative number of cases; red line corresponds to the best fit of the GLGM to the data. Grey lines correspond to the *m* bootstrapping realizations of the epidemic curves. Black lines correspond to the 95% CI bounds.

To model the second wave, we determine the epidemic profile by extending the entire uncertainty to a concrete period of time, adding to the early epidemic period, a forecast of 151 days ahead as described in section 3.4. In this case, the forecast starts at September 2, 2020 and finished at January 29, 2021. Figure 8 shows for the GLGM model, the best fit characterization of the first wave using *m* = 300 bootstrap realizations. In Figure 8.a, the blue circles are the daily cases reported. The solid grey lines correspond to the *m* bootstrapping realizations of the epidemic curves. The solid red line corresponds to the mean confidence or best fit of the GLGM respect the data; while solid black lines correspond to the upper and lower bounds of 95% confidence around the best fit of the model to the early epidemic. Also, it has been included in this figure, the curve of cumulative number of cases represented in Fig. 8.b as the solid blue line; the red line corresponds to the best fit of the GLGM to the data. Grey lines correspond to the m bootstrapping realizations of the epidemic curves. Black lines correspond to the 95% CI bounds. In both figures, it has been included a vertical slashed gray line to separate the calibration of the early epidemic period, to the forecasted period.

**Figure 8.**
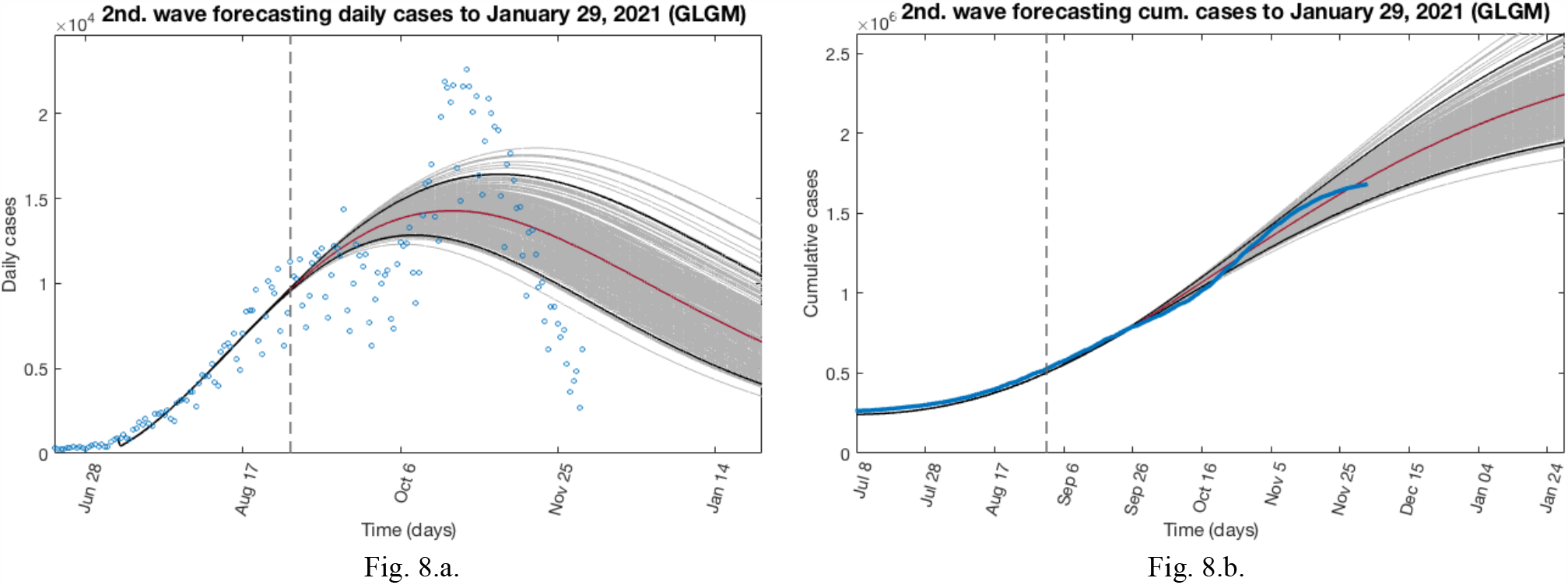
Characterization of the epidemic profile for the second wave using GLGM. In Fig. 8.a the blue circles are the cases reported daily; red line corresponds to the best fit of the GLGM to the data. Grey lines correspond to the *m* bootstrapping realizations of the epidemic curves. Black lines correspond to the 95% CI bounds. In Fig. 8.b, the solid blue line is the cumulative number of cases; red line corresponds to the best fit of the GLGM to the data. Grey lines correspond to the *m* bootstrapping realizations of the epidemic curves. Black lines correspond to the 95% CI bounds. The vertical slashed gray line to separate the calibration of the early epidemic period, to the forecasted period

## 5. Results and discussion

Simulation results and the main parameters that characterize both waves are shown in Table 1, bringing important information about the behavior of the epidemics over time. For the first wave, GGM and GLGM models indicate that its dynamics follows an exponential growth dynamics, with values of *p* ≈ 1, and the cumulative number of cases can be obtained by the equation *C* (*t*) = *C*_0_*e*^*rt*^. In the case of GLGM, the model estimates a final size of the epidemic *K* = 2.40·10^5^ (95% CI: 2.1·10^5^, 3.2·10^5^), whereas the data reported is 2.58·10^5^ cases This is displayed in Fig. 7.b. Also, the model is able to detect the peak of the epidemic for that first wave accurately.

**Table 1.** Main parameters that characterize the epidemic curves of the COVID-19 in Spain, from January 31, 2020 to a forecasted period to January 29, 2021.

| Model | Parameter | 1st. Epidemic Wave<br>(from January 31, 2020 to June 20, 2020) | 2nd. Epidemic Wave<br>(from July 8, 2020 to January 29, 2021) |
| --- | --- | --- | --- |
| GGM | $r$ | 0.29 (95% CI: 0.28, 0.30) | 3.6 (95% CI: 3.40, 3.70) |
| | $p$ | 0.96 (95% CI: 0.96, 0.97) | 0.64 (95% CI: 0.64, 0.65) |
| GLGM | $r$ | 0.24 (95% CI: 0.23, 0.25) | 6.8 (95% CI: 6.30, 7.30) |
| | $p$ | 1.0 (95% CI: 0.99, 1.0) | 0.59 (95% CI: 0.58, 0.60) |
| | $K$ | $2.48 \cdot 10^5$ (95% CI: $2.1 \cdot 10^5$, $3.2 \cdot 10^5$) | $2.48 \cdot 10^6$ (95% CI: $1.93 \cdot 10^6$, $3.24 \cdot 10^6$) |
| Reported | Cumulative Cases | $2.58 \cdot 10^5$ | $1.69 \cdot 10^6$ (as for December 3, 2020) |
| *SIR* | $\mathcal{R}_0 = S_0/\rho$ | 1.0025 | 1.0274 (estimated at January 29, 2021) |

In the case of the second wave, we have first estimate the epidemic profile to the last data available at the time to write this article. The GGM and GLGM models indicate that the epidemics curve follows a concave-down incidence pattern (0.5 < *p* < 1). In GGM, it has been obtained values of *p* = 0.64 (95% CI: 0.64, 0.65), which are near to 2/3, predicting that the incidence will grow quadratically, while the cumulative number of cases will follow a cubic polynomial pattern. Likewise, the GLGM model also indicates similar behavior for the epidemic growth, obtaining a value of *p* = 0.59 (95% CI: 0.58, 0.60). In this case, since the second wave is ongoing, the final size of the epidemic is forecasted. Hence, we have extended the entire uncertainty to January 29, 2021, obtaining *K* = 2.48·10^6^ (95% CI: 1.93·10^6^, 3.24·10^6^), when the current data indicate that as for December 3, 2020, the cumulative number of cases reported is 1,693,591. The profile of the cumulative number of cases, reported and forecasted, are shown in Fig. 8.a.

Overall, our analysis of empirical COVID-19 disease data in Spain, using GLGM model has revealed that, in the absence of an effective vaccine in the near months, the epidemic profile of the second wave can become in the third epidemic wave or, even more, it could remain in the population for long time, probably, to the end of summer 2021. This should be take into account in order to implement strict contention measures.

## 6. Conclusions and future work

In this paper, we have described the utilization of two phenomenological models to understand and characterize shifts in the epidemic growth patterns of the epidemics waves outbreaks of the COVID-19 in Spain. We have illustrated the proposed approach through simulations using data gathering of 44 weeks, and coupling the Generalized Growth Model (GGM) and the Generalized Logistic Growth Model (GLGM). Our findings indicate that even if the information about the disease is very limited or scattered, using an adequate combination of phenomenological models, it is possible to assess epidemic growth patterns over time, and even construct near or long terms forecasts.

In general, the model shows that in the absence of an effective vaccine, the second wave was likely inevitable and arrived just a few days after the end of the confinement. Likewise, the model shows that the delay in the implementation of strict control interventions at the beginning of this second wave, and the maintenance of these measures, may have caused an excess in the exponential growth of cases disease. This is reflected in the increase of the value of intrinsic growth rate *r* and, the basic reproductive number *ℛ*_0_ of the second wave, respect the first one. In this context, the basic reproductive number *ℛ*_0_ seems to be quite unrealistic, indicating that probably not all infected has been properly diagnosed and accounted. Hence, at the current state of the epidemic, it has been demonstrated that the strategy of adaptive or partial confinement may not be effective at all as a control measure, and even that a third wave is possible.

In future works, our goal is to provide a better approximation of the up to date epidemic curves of the COVID-19 in Spain, based on more complex phenomenological models. For this purpose, it is crucial to dispose of accurate and up to date sets of data series of diagnosed and reported cases.

## Data Availability

Data supporting the results reported in the published article can be found including, where applicable, hyperlinks to publicly archived datasets analysed or generated during the study.

https://datos.gob.es/es/catalogo/e05070101-evolucion-de-enfermedad-por-el-coronavirus-covid-19

